# Passive Monitoring of Physiological Data and Self-reported Symptoms to Detect Clusters of People with COVID-19

**DOI:** 10.1101/2020.07.06.20141333

**Authors:** Giorgio Quer, Jennifer M. Radin, Matteo Gadaleta, Katie Baca-Motes, Lauren Ariniello, Edward Ramos, Vik Kheterpal, Eric J. Topol, Steven R Steinhubl

## Abstract

Traditional screening for COVID-19 typically includes survey questions about symptoms, travel history, and sometimes temperature measurements. We explored whether longitudinal, personal sensor data can help identify subtle changes which may indicate an infection, such as COVID-19. To do this we developed an app that collects smartwatch and activity tracker data, as well as self-reported symptoms and diagnostic testing results from participants living in the US. We assessed whether symptoms and sensor data could differentiate COVID-19 positive versus negative cases in symptomatic individuals. Between March 25 and June 7, 2020, we enrolled 30,529 participants, of whom 3,811 reported symptoms, 54 reported testing positive for COVID-19, and 279 negative. We found that a combination of symptom and sensor data resulted in an AUC=0.80 [0.73 – 0.86] which was significantly better (p < 0.01) than a model which just considered symptoms alone (AUC=0.71 [0.63 – 0.79]) in the discrimination between symptomatic individuals positive or negative for COVID-19. Such orthogonal, continuous, passively captured data may be complementary to virus testing that is generally a one-off, or infrequent, sampling assay.

## INTRODUCTION

Due to the current lack of fast and reliable testing, one of the greatest challenges for preventing transmission of SARS-CoV-2 is the ability to quickly identify, trace, and isolate cases before they can further spread the infection to susceptible individuals. As regions across the U.S. start implementing measures to reopen businesses, schools, and other activities, many rely on current screening practices for COVID-19, which typically include a combination of symptom and travel-related survey questions and temperature measurements. However, this method is likely to miss pre-symptomatic or asymptomatic cases, which make up approximately 40% to 45% of those infected with SARS-CoV-2, and who can still be infectious.^1,2^ An elevated temperature (>100 degrees Fahrenheit) is not as common as frequently believed, being present in only 12% of individuals who tested positive for COVID-19,^3^ and just 31% of hospitalized COVID-19 patients at the time of admission.^4^

Smartwatches and activity trackers, which are now worn by 1 in 5 Americans,^5^ can improve our ability to objectively characterize each individual’s unique baseline for resting heart rate,^6^ sleep,^7^ and activity and therefore can be used to identify subtle changes in that users data which may indicate that they are coming down with a viral illness. Previous research from our group has shown that this method, when aggregated at the population level, can significantly improve real-time predictions for influenza-like illness.^8^ Consequently, we created a prospective app-based research platform, called DETECT (Digital Engagement & Tracking for Early Control, & Treatment), where individuals can share their sensor data, self-reported symptoms, diagnoses, and electronic health record data with the aim of improving our ability to identify and track individual and population level viral illnesses, including COVID-19.

A previously reported study that captured symptom data in over 18,000 SARS-CoV-2 tested individuals via a smartphone-based app found that symptoms were able to help distinguish between individuals with and without COVID-19.^2^ The aim of this study is to investigate if the addition of individual changes in sensor data to symptom data can be used to improve our ability to identify COVID-19 positive versus COVID-19 negative cases among participants who self-reported symptoms.

## METHODS

### Study population

Any person living in the United States over the age of 18 years old is eligible to participate in the DETECT study by downloading the iOS or Android research app, MyDataHelps. After consenting into the study, participants are asked to share their personal device data (including historical data collected prior to enrollment), report symptoms and diagnostic test results, and connect their electronic health records. Participants can opt to share as much or as little data as they would like. Data can be pulled in via direct API with Fitbit devices, and any device connected through Apple HealthKit or GoogleFit data aggregators. Participants were recruited via the study website (www.detectstudy.org), media reports, and outreach from our partners at Fitbit, Walgreens, CVS/Aetna, and others.

Between March 25, 2020 and June 7, 2020, our research study enrolled 30,529 individuals with representation from every state in the United States. Among the consented individuals, 62.0% are female and 12.8% are 65 or more years old. 78.4% of participants connected their Fitbit device to the study-app, 31.2% connected the data from Apple Health Kit while 8.1% connected data from Google Fit (note that one individual can connect to multiple platforms). In addition, 3,811 reported at least one symptom (12.5%), and of those 54 also reported testing positive for COVID-19, and 279 reported testing negative for COVID-19. The number of days per different data types and data aggregator system is presented in Table 1, while the symptoms distribution for symptomatic individuals tested for COVID-19, or not tested is shown in Figure 1.

**Table 1:** Participants characteristics and device usage.

|  | Total | Any symptom | COVID-19 positive | COVID-19 negative | P-value |
| --- | --- | --- | --- | --- | --- |
| **Demographic Data** |  |  |  |  |  |
| **Number of participants** | 30529 | 3811 | 54 | 279 | - |
| **Female [%]** | 62.0% | 74.2% | 79.6% | 71.3% | 0.65 |
| **Over 65 [%]** | 12.8% | 5.7% | 1.9% | 7.5% | 0.22 |
| **Fitbit Users [%]** | 78.4% | 86.7% | 85.2% | 84.9% | 1.00 |
| **Apple Users [%]** | 31.2% | 32.0% | 38.9% | 33.7% | 0.66 |
| **Sensor Data** |  |  |  |  |  |
| **Available days (IQR)** |  |  |  |  |  |
| **RHR** | 322<br>(131-387) | 325<br>(153-396) | 312<br>(98-377) | 300<br>(119-392) | 0.70 |
| **Sleep** | 249<br>(48-373) | 273<br>(105-383) | 283<br>(52-362) | 246<br>(70-375) | 0.56 |
| **Activity** | 394<br>(370-412) | 407<br>(379-415) | 404<br>(375-410) | 401<br>(374-413) | 0.57 |
| **Mean Change (SD)** |  |  |  |  |  |
| **RHR (bpm)** | ~ 0<br>(2.84 ) | 0.40<br>(3.18 ) | 1.15<br>(4.83 ) | 0.61<br>(3.68 ) | 0.33 |
| **Sleep (min)** | ~ 0<br>(54 ) | 3<br>(59 ) | 57<br>(92 ) | 4<br>(68 ) | < 0.01 |
| **Activity (steps)** | 52<br>(2659 ) | -323<br>(2771 ) | -3533<br>(4418 ) | -208<br>(3086 ) | < 0.01 |

**Figure 1:**
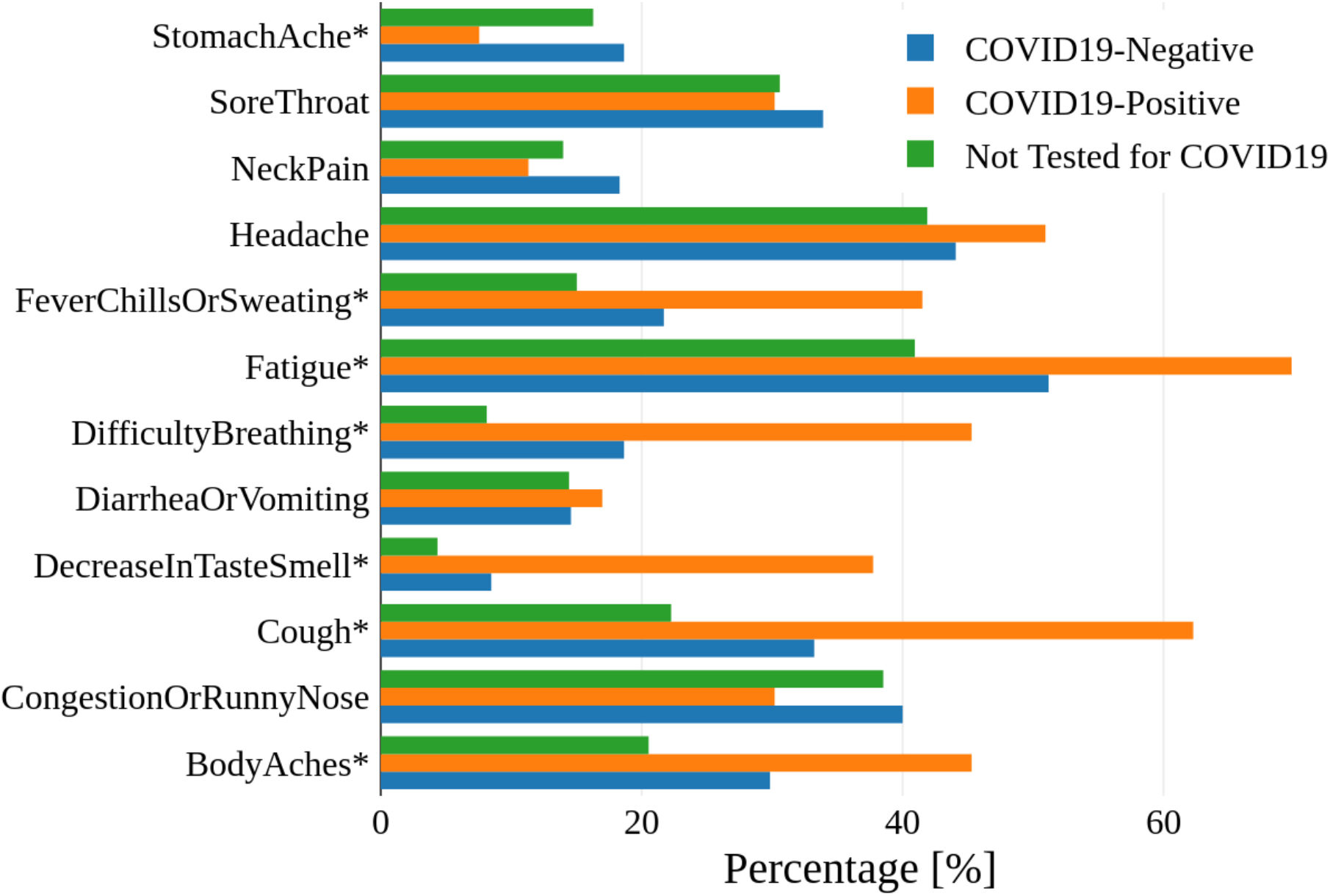
Frequency of symptoms among participants. Participants who reported at least one symptom have been divided into 3 cohorts: participants negative and positive to a COVID-19 test, and participants who did not undergo a test for COVID-19. The frequency of each specific symptom is reported in the figures for the three cohorts. Symptoms with significant difference between COVID-19 positive and negative participants (p-value < 0.05 of Fisher’s exact test) are marked with an asterisk in the figure.

### Ethical Considerations

The protocol for this study was reviewed and approved by the Scripps Office for the Protection of Research Subjects. All individuals participating in the study provided informed consent electronically.

### Statistical Analysis

Only participants with self-reported symptoms and COVID-19 test results were considered in this analysis. For each participant, two sets of data were extracted: the *baseline data*, which included signals spanning from 21 to 7 days before the reported start date of symptoms, and the *test data*, which included signals beginning the first date of symptoms to 7 days after symptoms. Three types of data were considered from personal sensors: daily resting heart rate (*DailyRHR*), sleep duration in minutes (*DailySleep*) and activity based on daily total step count (*DailyActivity*). The daily resting heart rate is calculated by the specific device.^35^ The total amount of sleep for a given day was based on the total period of sleep between 12 noon of the current day to 12 noon of the next day. When multiple devices from the same individual provide the same information, Fitbit device data was prioritized for consistency. Overlapping data were combined minute by minute, before aggregating for the whole day.

A single baseline value per individual was extracted for each data type by considering the median value over the individual’s baseline data. This value is representative of a participant’s “normal” before the reported symptoms. The baseline value was compared to the test data as follows:

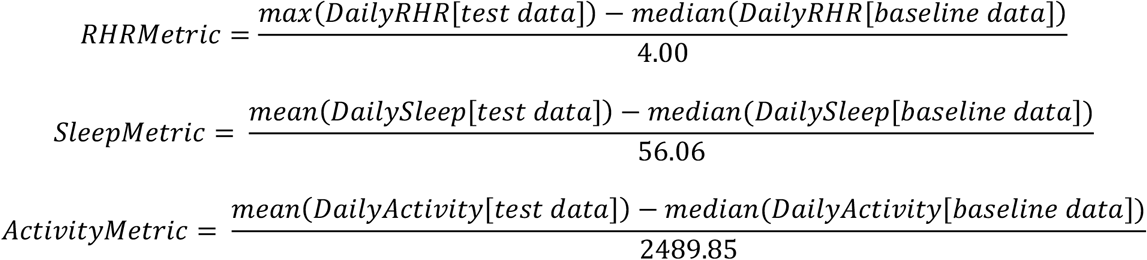

Values were normalized to have a unitary interquartile range using normalization parameters calculated on all data recorded. For all these metrics, values close to zero indicate small variations from baseline values.

For the metric based on symptoms only, we adapted the results from the study by Menni et al.^2^ to our available data:

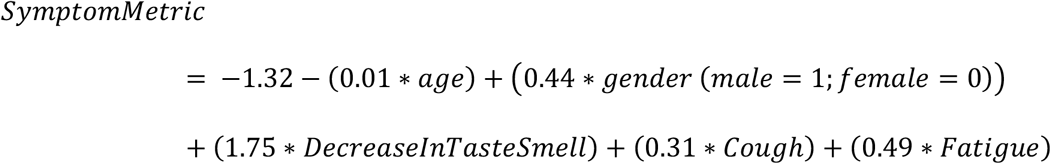

A simple manual metric aggregation strategy without optimization was used to enable a clear understanding of the benefits provided when data from multiple sources were considered together. The aggregated metrics were:

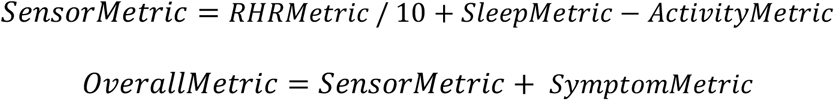

The main outcomes are receiver operating characteristic (ROC) curves for each of the proposed metrics. The curves are obtained by considering a binary classification task between participants self-reported as COVID-19 positive and negative. Confidence intervals, reported with a confidence level of 95%, are estimated using bootstrap method by repeatedly sampling the dataset with replacement. The sampling is performed in a stratified manner, i.e., the balance of the classes is maintained over all experiments. Values for sensitivity (SE), specificity (SP), positive predictive value (PPV) and negative predictive value (NPV) were also calculated (Figure 2). These values are based on the point in the ROC with the optimal trade-off between sensitivity and specificity, which may vary depending on the shape of the curve. For each metric analyzed, we applied the Mann-Whitney U test to investigate the statistical difference among positive and negative class values and we reported the p-value. The comparison metric to assess the overall performance was the area under curve (AUC) of the ROC.

**Figure 2:**
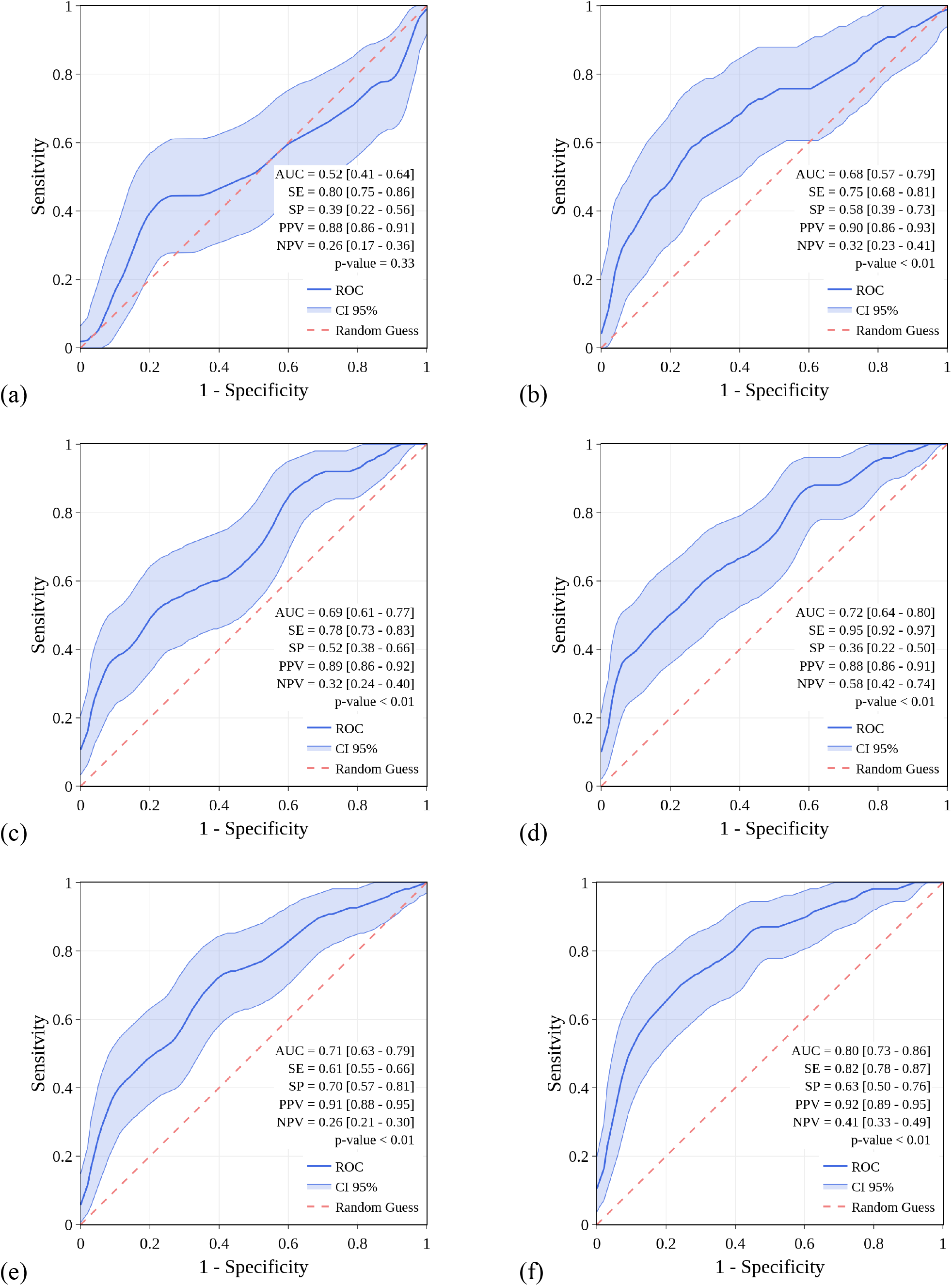
Prediction of COVID-19 from self-reported symptoms and sensor data. ROC for the discrimination between COVID-19 positive and COVID-19 negative based on the available data: resting heart rate data (a); sleep data (b); activity data (c); all available sensor data (d); symptoms only (e); and symptoms with sensor data (f).

## RESULTS

The symptoms distribution for symptomatic individuals tested for COVID-19, or not tested is shown in Figure 1.

### Sensor Data

A minority of symptomatic participants (30.3%) who tested for COVID-19 had an RHR greater than 2 standard deviations above the average baseline value during symptoms. Change in RHR on its own (Table 1) did not allow for significant discrimination between COVID-19 positive and negative subjects using the RHRMetric (AUC of 0.52 [0.41 – 0.64]). (Figure 2a) Sleep and activity did show a significant difference among the two groups, (Table 1) with an AUC of 0.68 [0.57 – 0.79] for the SleepMetric (Figure 2.b) and 0.69 [0.61 – 0.77] for the ActivityMetric (Figure 2.c), supporting that the sleep and activity of COVID-19 positive participants were impacted significantly more than COVID-19 negative participants. Sleep and activity are slightly correlated, with a negative correlation coefficient of −0.28, p-value < 0.01.

To evaluate the contribution of all the data type commonly available through personal devices, we combined the RHR, sleep, and activity metrics in a single metric (SensorMetric, Figure 2.d). This improved the overall performance from the three sensor metrics to an AUC of 0.72 [0.64 – 0.80].

### Symptom Data

We also considered a model only based on self-reported symptoms (SymptomMetric, Figure 2.e), along with age and sex. With respect to the previously published model,^2^ we measure a slightly lower AUC of 0.71 [0.63 – 0.79].

### Combined Symptoms & Sensor Data

When participant-reported symptoms and sensor metrics are jointly considered in the analysis (OverallMetric, Figure 2.f), the achieved performance was significantly improved (p < 0.01), relative to either alone, with an AUC of 0.80 [0.73 – 0.86].

## DISCUSSION

Our results show that individual changes in physiologic measures captured by most smartwatches and activity trackers are able to significantly improve the distinction between symptomatic individuals with and without a diagnosis of COVID-19 beyond just symptoms alone. While encouraging, these results are based on a relatively small sample of participants. This work builds on our earlier retrospective analysis demonstrating the potential for consumer sensors to identify individuals with influenza-like illness, which has subsequently been replicated in a similar analysis of over 1.3 million wearable users in China for predicting COVID-19.^8,9^ In response to the COVID-19 pandemic a number of prospective studies, led by device manufacturers and/or academic institutions, including DETECT, accelerated deployment to allow interested individuals to voluntarily share their sensor and clinical data to help address the global crisis.^10-14^ The largest of these efforts, Corona-Datenspende, was developed by the Robert Koch Institut in Germany and has enrolled over 500,000 volunteers.^15^

As different individuals experience a wide range of symptomatic and biologic responses to infection with SARS-CoV-2, it is likely that their measurable physiologic changes will also vary.^16-18^ For that reason, it is possible that biometric changes may be more valuable in identifying those at highest risk for decompensation rather than just a dichotomous distinction in infection status. Due to limited testing in the United States, especially early in the spread of the COVID-19 pandemic, individuals with more severe symptoms may have been more likely to be tested. Consequently, the ability to differentiate between COVID-19 positive and negative cases based on symptoms and sensor data may change over time as testing increases, and as other upper respiratory illnesses such as seasonal influenza increase this fall.

The early identification of symptomatic and pre-symptomatic infected individuals would be especially valuable as transmission is common and people may potentially be even more infectious during this period.^19-21^ Even when individuals have no symptoms, there is evidence that the majority have lung injury by CT scan, and a large number have abnormalities in inflammatory markers, blood cell counts and liver enzymes.^18,22-24^ As the depth and diversity of data types from personal sensors continues to expand—such as heart rate variability (HRV), respiratory rate, temperature, oxygen saturation, and even continuous blood pressure, cardiac output and systemic vascular resistance—the ability to detect subtle individual changes in response to early infectious insults will potentially improve and enable the identification of individuals without symptoms.

In the past, the normality of a specific biometric parameter, such as resting heart rate, duration of nightly sleep, and daily activity, was based on population norms. For example, a normal RHR is generally considered anything between ∼60-100 BPM. However, recent work looking at individual daily RHRs over two years found that each person has a relatively consistent RHR, for them, that fluctuates by a median of only 3 BPM weekly.^6^ On the other hand, what would be considered normal RHR for an individual can vary by as much as 70 BPM (between 40 and 109 BPM) between individuals. The potential value in identifying important changes in an individual’s RHR as an early marker for COVID-19 infection is suggested by the description of 5,700 hospitalized COVID-19 patients.^4^ At the time of admission, a greater percentage of individuals had a heart rate of >100 BPM (43.1%) than had a fever (30.7%). Similarly, work in primate models of other viral and bacterial infections found that a significant increase in heart rate can be detected ∼2 days prior to a fever. ^25^

Just as individuals have heart rate patterns that are unique to them, the same is true for sleep patterns. While population norms for sleep duration have been defined by one-time survey data,^26^ longitudinal analysis of daily sleep over several years support much greater variation in what is normal for a specific individual.^7^ Recognizing what is normal for an individual enables much earlier detection of deviations from that normal.

A strategy of test, trace and isolate has played a central role in helping control the spread of COVID-19. However, testing comes with many challenges including the enormous logistical and cost hurdles of recurrently testing asymptomatic individuals. In addition, testing in a population with very low prevalence can lead to a high proportion of false positive cases. A refined predictive model, based on personal sensors, could potentially enable an early, individualized testing strategy to improve performance and lower costs. Early testing may make the use of a contact tracing app more effective by identifying positive cases in advance and allowing for early isolation.

DETECT, and similar studies, also represents the transitioning of research from a dependence on brick and mortar research centers to a remote, direct-to-participant approach now possible through a range of digital technologies, including an ever-expanding collection of sensors, applications of machine learning to massive data sets, and the ubiquitous connectivity that enables rapid 2-way communications 24/7.^27,28^ The promise of digital technologies is that their evolution will continue to bring us closer to identifying the best combination of measures and associated algorithms that identify infection with SARS-CoV-2 or other pathogens. However, it is equally critical to develop and continuously improve on an engaging digital platform that provides value to participants and researchers. This has proven to be extremely challenging with a recent analysis of 8 different digital research programs involving 100,000 participants have a median duration of retention of only 5.5 days.^29^ Digital trials such as DETECT also do come with unique challenges to assure privacy and security, which can only be dealt with by effectively informing participants before consent, storing the data with the needed level of security and providing access to the data only for research purposes.^30^ App-based contact tracing, which is not part of DETECT, is an especially sensitive and ethically complicated use of digital technology that can be used to address the pandemic.^31^

### Limitations

Our analyses are dependent entirely on participant-reported symptoms and testing results, as well as the biometric data from their personal devices. Although this is not consistent with the historically more common direct collection of information in a controlled lab setting or via electronic health records, previous work has confirmed their value and their accuracy beyond data routinely captured during routine care.^32-34^ Additionally, individuals owning a smartwatch or activity tracker and having access to COVID-19 diagnostic testing may not be fully representative of the general population. Finally, in the early version of the DETECT app we were not able to track the duration or trajectory of individual symptoms, care received and eventual outcomes.

## Conclusion

These preliminary results suggest that sensor data can incrementally improve symptom-only based models to differentiate between COVID-19 positive and negative symptomatic individuals, which has the potential to enhance our ability to identify a cluster before more spread occurs. Such orthogonal, continuous, passively captured data may be complementary to virus testing that is generally a one-off, or infrequent, sampling assay.

## Data Availability

This is an ongoing study. We plan to make the de-identified data widely available, we are currently working on a procedure to do it.

## AUTHORS CONTRIBUTIONS

G.Q, J.M.R., K.B., V.K., E.J.T., and S.R.S. made substantial contributions to the study conception and design. K.B., L.A., E.R., V.K., S.R.S. made substantial contributions to the acquisition of data. G.Q. and M.G. conducted statistical analysis. G.Q, J.M.R., M.G. and S.R.S. made substantial contributions to the interpretation of data. G.Q., J.M. R. and S.R.S. drafted the first version of the manuscript. G.Q, J.M.R., M.G., K.B., L.A., E.R., V.K., E.J.T., and S.R.S. contributed to critical revisions and approved the final version of the manuscript. G.Q., J.M.R., and S.R.S. take responsibility for the integrity of the work.

## COMPETING INTERESTS

S.R.S. reports grants from Janssen and personal fees from Otsuka and Livongo, outside the submitted work. G.Q., J.M.R., M.G., K.B., L.A., E.R., V.K. and E.J.T. report no competing interests.

Summary of the collected data and demographic information about the cohort. Available days are specified for each data type, with median and interquartile range (IQR) values. Changes in RHR, Steps, and Sleep from baseline (−21 to 7 days) to symptomatic period (0-7 days) are reported, where for individuals with no symptoms we consider March 6, 2020 as the day 0. p-values are evaluated comparing COVID-19 positive and negative groups.

